# Artificial Intelligence Algorithms in Nailfold Capillaroscopy Image Analysis: A Systematic Review

**DOI:** 10.1101/2024.07.28.24311154

**Authors:** Omar S. Emam, Mona Ebadi Jalal, Begonya Garcia-Zapirain, Adel S. Elmaghraby

## Abstract

**Background:** Non-invasive imaging modalities offer a great deal of clinically significant information that aid in the diagnosis of various medical conditions. Coupled with the never-before-seen capabilities of Artificial Intelligence (AI), uncharted territories that offer novel innovative diagnostics are reached. This systematic review compiled all studies that utilized AI in Nailfold Capillaroscopy as a future diagnostic tool.

**Methods and Findings:** Five databases for medical publications were searched using the keywords artificial intelligence, machine learning, deep learning and nailfold capillaroscopy to return 105 studies. After applying the eligibility criteria, 10 studies were selected for the final analysis. Data was extracted into tables that addressed population characteristics, AI model development and nature and results of their respective performance. We found supervised deep learning approaches to be the most commonly used (*n* = 8). Systemic Sclerosis was the most commonly studied disease (*n* = 6). Sample size ranged from 17,126 images obtained from 289 participants to 50 images from 50 participants. Ground truth was determined either by experts labelling (*n* = 6) or known clinical status (*n* = 4). Significant variation was noticed in model training, testing and feature extraction, and therefore the reporting of model performance. Recall, precision and Area Under the Curve were the most used metrics to report model performance. Execution times ranged from 0.064 to 120 seconds per image. Only two models offered future predictions besides the diagnostic output.

**Conclusions:** AI has demonstrated a truly remarkable potential in the interpretation of Nailfold Capillaroscopy by providing physicians with an intelligent decision-supportive tool for improved diagnostics and prediction. With more validation studies, this potential can be translated to daily clinical practice.

## 1 INTRODUCTION

Capillaries are the tiniest and most numerous blood vessels that link the body’s arterial system to its venous system. They branch superficially and deeply into all body tissues to provide nutrients and remove waste products (1, 2). This healthy microenvironment can get severely dysfunctional due to the pathological processes in numerous abnormal conditions (3); for example: a) systemic diseases such as diabetes (4, 5), or metabolic syndrome, b) auto-immune inflammatory pathologies such as vasculitis and dermatomyositis (6), and c) connective tissue diseases (7) such as systemic sclerosis (SSc) (8, 9) systemic lupus erythematosus (10, 11), and Raynaud’s phenomenon (12, 13). Capillaries in the retina, tongue, or nailfolds provoke a particular medical interest due to their ease of accessibility and examination using common non-invasive tools that yield clinically significant information. In other words, the diagnosis and follow up of internal systemic conditions can be performed without the need to resort to invasive approaches.

Nailfold Capillaroscopy (NFC), which is a type of microscopic angioscopy, is a technique of visualizing the capillaries in the nailfold area (14). It examines the ultimate capillary endings in the finger as capillaries loop to turn back around. To visualize these terminal capillaries in that thin layered area of the skin, the subject as well as the surrounding environment are prepared and then a microscopic lens can be used to directly observe the capillaries. Nailfold Video Capillaroscopy (NVC) (15, 16)uses a more advanced scope with a camera that offers far superior resolution, clarity, and the ability to record a video and take photos (6). These images are then analyzed by trained experts to differentiate normal healthy capillaries from faulty pathological ones according to certain specific criteria. This information can aid in diagnosis, progression and severity assessment (17), disease staging (18–22), follow-up and perhaps prediction of certain medical conditions. Unfortunately, such convenient classical methods of manual analyses imply subjectivity, prolonged analyses time, and ambiguity of findings (23–25).

The recent trends utilizing Artificial Intelligence (AI), especially in medicine and biomedical sciences, seem to offer a highly sought-after outturn as a superior alternative (26, 27). To illustrate, many deep learning models have demonstrated the ability to objectively analyze images from NFC/NVC with higher, or at least comparable, efficiency as capillaroscopy experts in a significantly lower time (5, 15, 28–41). Advances in computer vision algorithms allow the extraction, quantification, and accurate analysis of far more features compared to human experts. These innovations present an unprecedented potential to link a multitude of variables to diseases and, consequently, draw future predictions (7). The information gained from such technology would be crucial to inform decisions concerning patient education and treatment in many ways. For example, predictive models can be used as an adjunct screening tool, diagnostic tools can help accelerate clinical work, establish risk stratification, and reduce rates of misdiagnoses (42). Moreover, the smart ‘learning’ nature of these tools will benefit from feedback and provide explanations to further improve their accuracy (43). As such, these innovations claim the potential to transform existing medical practices by enhancing the decision-making process using such tools. In other words, is it practically feasible to improve the quality of care delivered to patients using such cheap, easy, non-time-consuming means? In this systematic review, we aim to give an overview on the state-of-the-art by compiling all studies, to date, that utilized an AI algorithm to analyze output from NFC/NVC as a tool to be used clinicians in medical practice. We describe the methodology used to conduct a comprehensive search of the literature to encompass all novel studies addressing this topic. Next, we presented the summarized results of the included reports. Finally, we divided the discussion into four sections to organize the gathered evidence to answer our research questions with commentary on a few important issues as follows: Section I: a brief background on the significance of NFC in medicine, Section II: Challenges facing the manual method, Section III: How AI is solving these challenges, and finally Section IV: Future directions and limitations.

## 2 METHODS

This systematic review was conducted in compliance with the Preferred Reporting Items for Systematic Reviews and Meta-Analysis (PRISMA) (44). We formulated the research questions, and explained their significance, in **Table 1**, to guide the following search and screening steps.

**Table 1.** Research questions and their rationale.

|  | Research Question | Rationale |
| --- | --- | --- |
| Q1 | Can AI successfully detect clinically significant changes in NFC images? | To ensure that the information captured is relevant and useful to clinicians |
| Q2 | Is pre-analysis processing required for an optimized output? | To check the user-friendliness and practically consider the time elapsed |
| Q3 | Is there a minimum number of variables/features to be extracted for an accurate enough output? | To understand how these algorithms work and study the “significant” variables/features |
| Q4 | Is information from NFC/NVC alone enough? Or is combining them with other diagnostic modalities and data sources needed? | To maximize the potential for detection, data acquisition and interpretation |
| Q5 | Would the “early” use of such technologies help in predicting future outcomes? | To consider the possibility of early interventions and thus better patient reported outcomes |
| Q6 | What conditions/diseases can this technology be applied to? | To understand the scope and generalizability or individuality of these AI applications |

### 2.1 Data Collection

#### 2.1.1 Search Strategy

We performed an all-time search on December 14^th^, 2023, that was later updated in March 2024, utilizing five electronic medical databases: PubMed (including MEDLINE), EMBASE, CINAHL, and Web of Science using the keywords: “*Nailfold*”, “*Capillaroscopy*”, *“Artificial Intelligence”*, “*Machine Learning*” and “*Deep Learning*” to generate the search string*: (nail OR nailfold OR “nail-fold”) AND (capillaroscopy OR “video capillaroscopy” OR NVC OR “microscopic angioscopy” OR onychoscopy) AND (“artificial intelligence” OR AI OR “machine learning” OR “deep learning” OR algorithm*).* MeSH terms and boolean operators were used where appropriate. **Fig.1** shows a PRISMA flow diagram that summarizes the searching process.

**Fig. 1.**
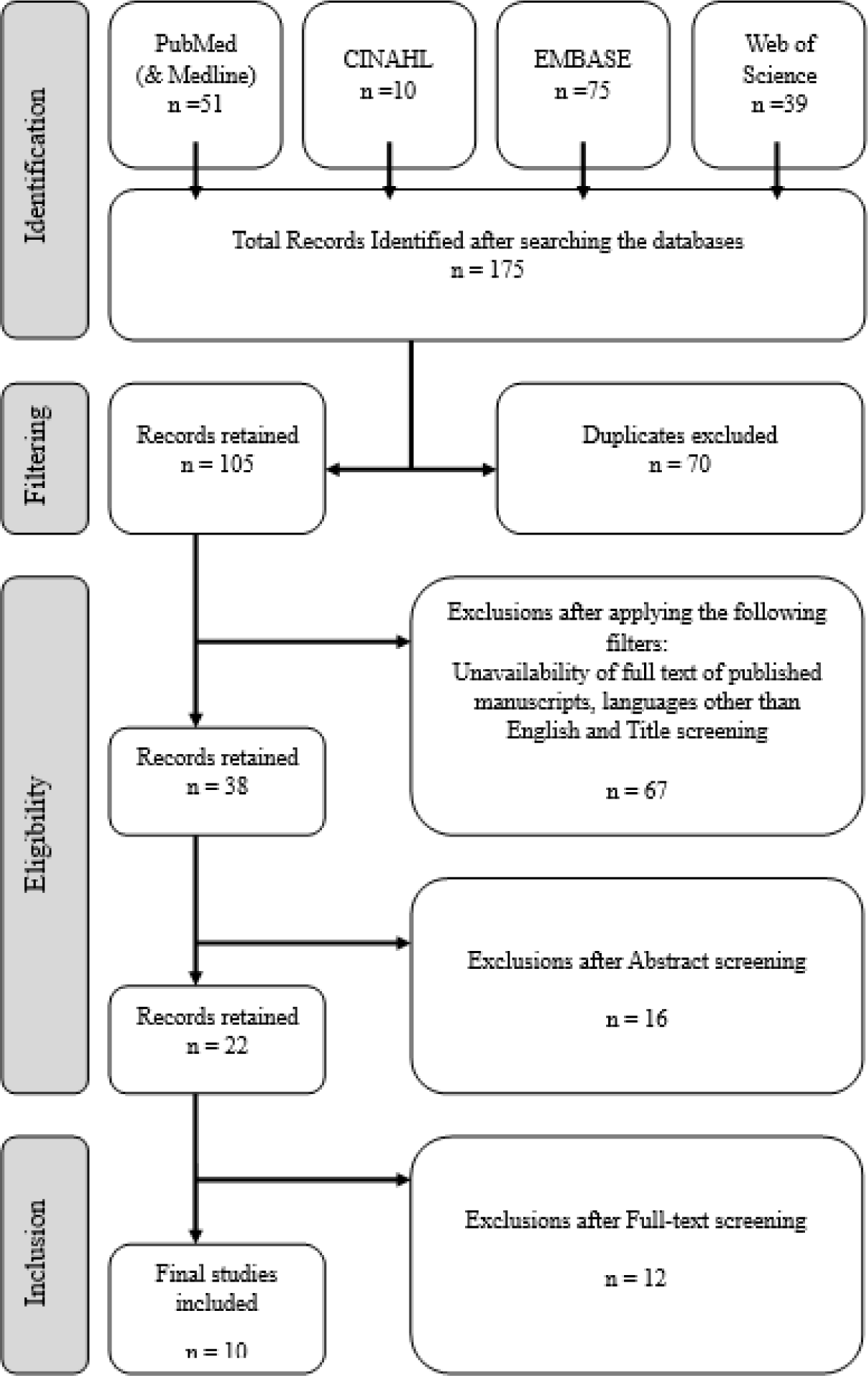
PRISMA Flow diagram summarizing the search and screening process.

#### 2.1.2 Inclusion and Exclusion Criteria

We limited the search results to availability of full texts in the English language through our institutional access. We included all 1) peer reviewed journal articles, 2) both qualitative and quantitative studies, 3) that presented an AI model, 4) to analyze NFC/NVC images, and 5) was internally validated or tested on a real dataset.

We excluded unpublished data under review, conference abstracts and proceedings, and grey literature such as short surveys, and letters to editors. Additionally, we excluded studies that 1) utilized a purely mathematical model/algorithm that is not categorized as AI, or 2) presented technical aspects of the technology without applying it to a real data set as a diagnostic or prognostic tool, or 3) included other techniques to study the nailfold capillaries other than NFC such as optoacoustic imaging or ultrasound, whether independently or in fusion with NFC.

#### 2.1.3 Study Selection and Data Extraction Process

Two authors independently performed the search and imported the results to the EndNote v.20 reference manager, where duplicates were removed. Any conflicting judgements were resolved by a third author. We then extracted data related to the study design, target population characteristics and sampling, pre-procedural setting description, pre-analysis processing, AI methods, model training and development, type of input and output, performance metrics, limitations, strengths, and finally conclusions; that process was repeated for all final ten studies included in the review.

### 2.2 Quality Assessment and Risk of Bias

Assessment of quality and the risk of bias in diagnostic accuracy studies is commonly done using known validated tools such as STARD (45) and QUADAS (46), or tools like TRIPOD (47) for prognostic studies as well. However, the novel nature of AI-related studies created a demand for more relevant and appropriately updated tools that many authors sought to meet by modifying or adding extensions to the aforementioned tools such as QUADAS-AI (48), STARD-AI (49), and TRIPOD-AI (50). Unfortunately, these tools are still under development analysis and there is currently no agreed upon gold standard tool to be used (51). For these reasons we opted to refer to the updated QUADAS-2 tool (52) and complement the assessment with the MINIMAR (53) and CAIR (54) tools that present a checklist for reporting AI studies to healthcare providers, as outlined in supplementary document **S1**.

## 3 RESULTS

The search process returned 105 results that were then screened by title, abstract, and finally after full-text reading using the eligibility criteria to arrive at the final list of ten studies as summarized in **Fig. 1** (15, 28–32, 35, 40, 41, 55). Six out of the ten studies were published within a year of writing this manuscript (in 2023) illustrating the novelty of the topic at hand (28–31, 41, 55). The geographical distribution of countries where these studies were conducted is shown in **Fig. 1**. The following **Table *2*** shows a summary of the main highlights of all ten studies including population characteristics.

**Fig. 1.**
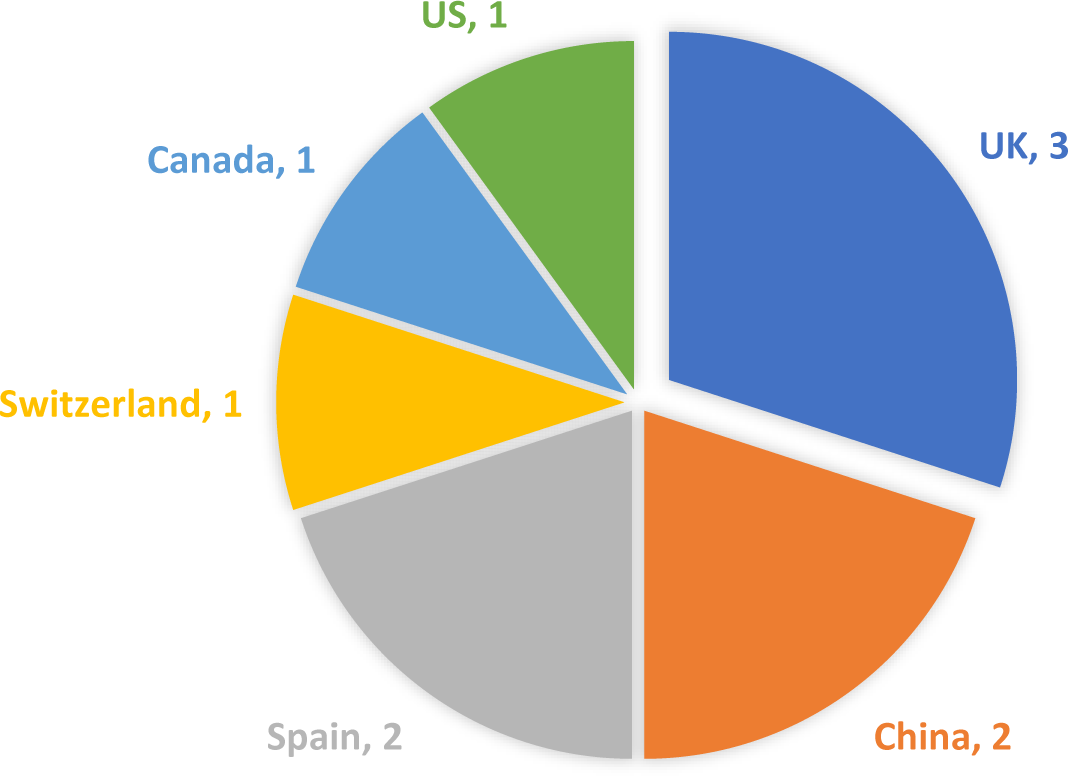
Geographical distribution of the countries of included studies.

**Table 1.**
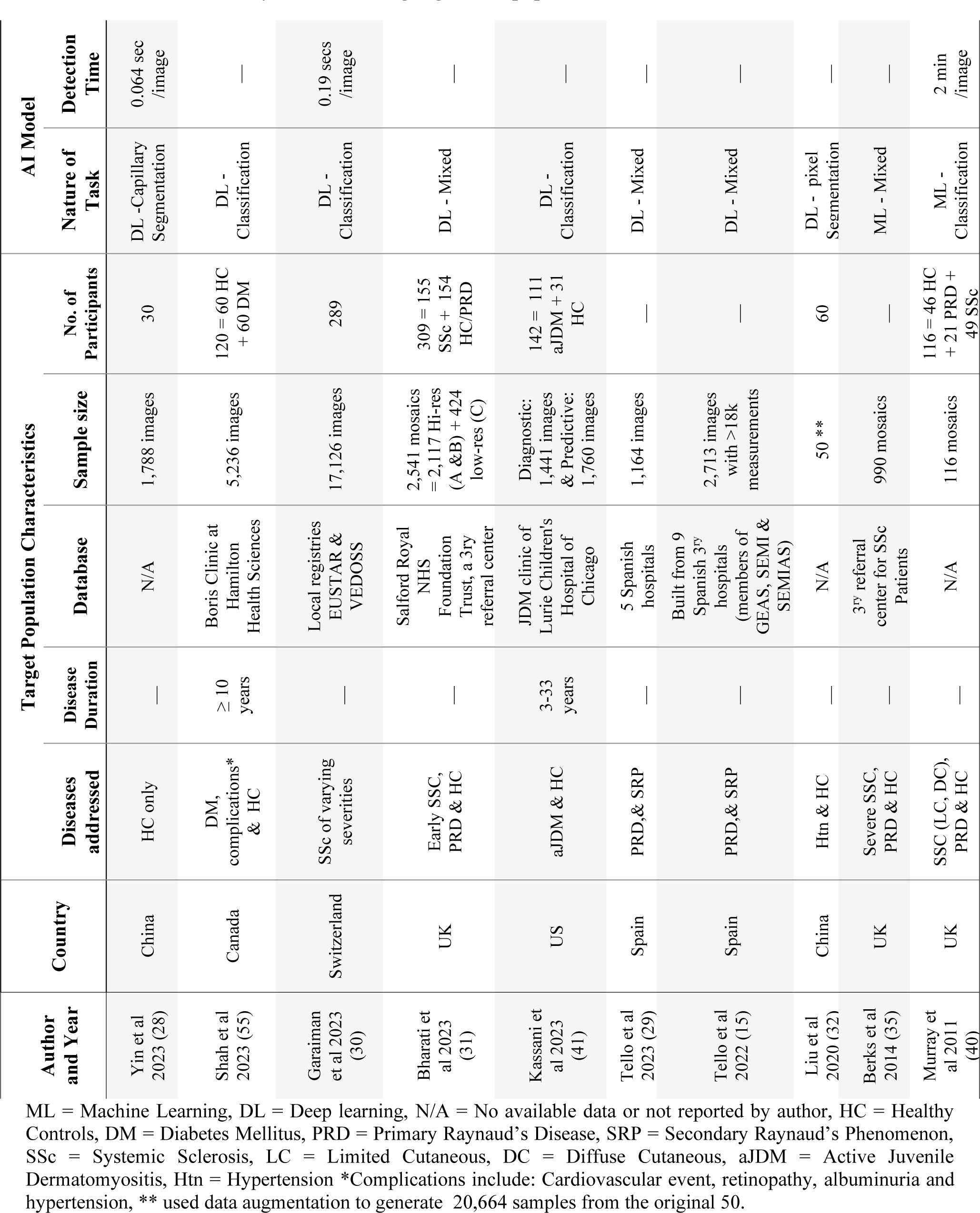
A summary of the main highlights and population characteristics of all ten studies.

## Synthesis of Evidence

### 3.1 Study Designs

Consistent with most diagnostic accuracy studies, all ten investigators adopted a non-experimental cross-sectional design with case-control selection (15, 28–32, 35, 40, 41, 55). After recruiting participants via certain criteria, the developed AI algorithm was tested against the ground truth to assess the model’s performance.

### 3.2 Population Characteristics

Yin et al was the only author to include healthy volunteers as the sole subject of his study (28). Alternatively, Garaiman et al was the only investigator to acknowledge limiting his study to include only diseased patients due to ethical consent considerations (30). The remaining eight studies included both diseased participants and normal controls (15, 29, 31, 32, 35, 40, 41, 55). Without including both normal and diseased subjects, an AI algorithm would not be capable of predicting abnormalities. The diseases addressed by each study are shown in **Fig. *2***.

**Fig. 2.**
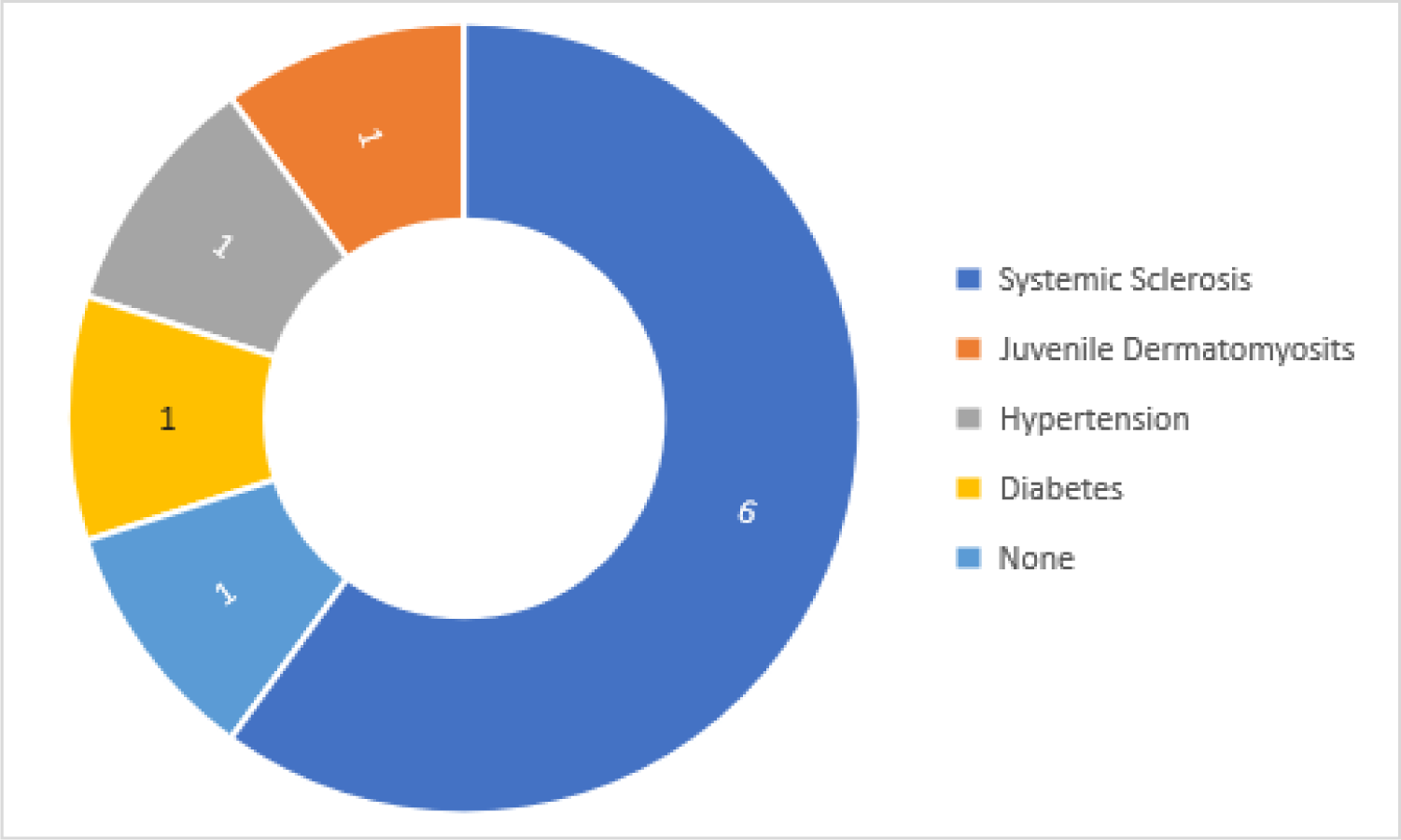
Diseases addressed by the included studies.

Among the six studies investigating systemic sclerosis (SSc), three authors considered two variants as the normal controls: completely disease-free healthy participants and others with a common benign condition known as primary Raynaud’s phenomenon (31, 35, 40); also known as Raynaud’s disease (PRD) (56). Conversely, Garaiman et al and Tello et all were satisfied with PRD only as normal controls (15, 29, 30). Interestingly, despite the recognized significance of disease duration due to its pathological impact on the body, and, consequently, the outcomes assessed by NFC, only three authors (40, 41, 55) reported the disease duration of their respective participants.

Participants’ demographics were surprisingly not addressed in 50% of the studies (15, 28, 29, 32, 35). Moreover, only Kassani et al and Shah et al reported racial backgrounds, and thus, shedding light on a very significant - yet overlooked – factor: skin tone (41, 55). This finding highlights a significant gap in addressing imbalances due to a particularly relevant factor like skin-color, in addition to generally important variables such as sex, age, and other co-morbidities, that may influence the algorithm’s learning and output. Among all studies that investigated adults, Kassani et al was also unique in investigating a condition prevalent in the children’s age group, juvenile dermatomyositis (JDM) (41).

### 3.3 Sample Sizes

The highest sample size was 17,126 images obtained from 289 participants, as reported by Garaiman et al (30); which was more than three times the second highest sample size of 5,236 from 120 participants reported by Shah et al (55). Liu et al developed his algorithm with the least sample size of 50 images (32). **Fig. 3**Error! Reference source not found. shows a comparison of the total sample sizes of both images and participants across all included studies.

**Fig. 3.**
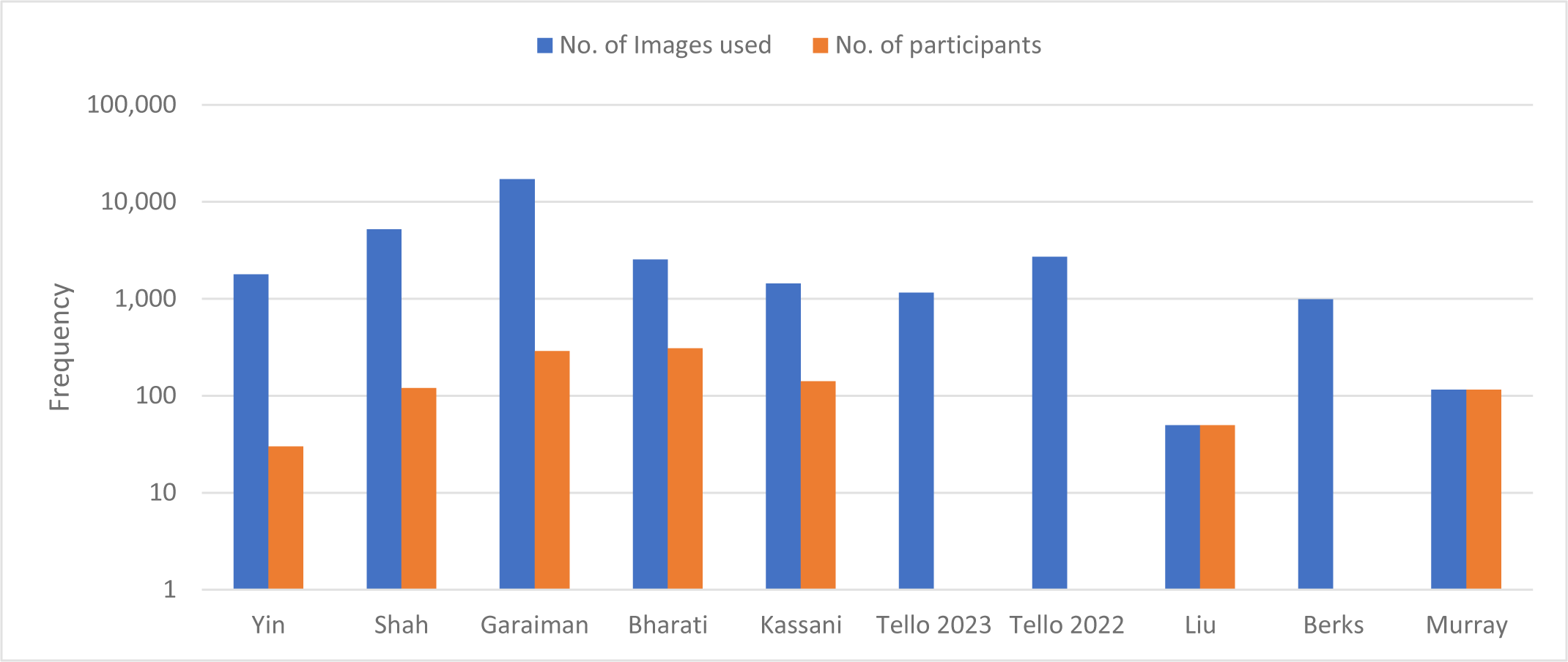
Comparison of the total sample sizes across all included studies shown in logarithmic scale.

Authors differed in their approach to obtaining sample images from participants, and in how balanced the normal controls and main study groups were. For instance, five authors obtained multiple images from multiple sections of multiple nailfolds spanning multiple encounters (28, 30, 31, 41, 55). Conversely, both Liu et al and Murray et al obtained a single image per participant (32, 40). However, Murray’s image is a mosaic, which is a panoramic wholesome image of the entire nailfold, compared to Liu’s single image that represents a section of the nailfold. Nonetheless, Liu et al used a data augmentation technique to generate a much bigger sample size, derived from the original 30, that reached 20,664 images (32). Alternatively, Tello et al chose images at random from a bigger pool of a previously prepared dataset without reporting the number of participants (15, 29). Similarly, Berks et al used 990 mosaics without reporting the number of fingers, participants, or encounters that contributed to that dataset (35). Finally, Kassani et al had a sample of 1,441 images for their diagnosis model and a different sample of 1,760 images for the predictive model (41).

### 3.4 AI Algorithms Development

#### 3.4.1 Model Architecture

The majority of authors employed a deep learning model to accomplish their task (15, 28–32, 41, 55), except for Berks et al and Murray et al who utilized a pipeline of machine learning models (35, 40). All ten authors utilized a supervised machine learning approach where the input images were labelled with the ground truth so that the model learns through the input-output pairs.

**Table *3*** summarizes the different architectures for each algorithm, learning approaches, diagnostic models’ development and performance.

**Table 3.**
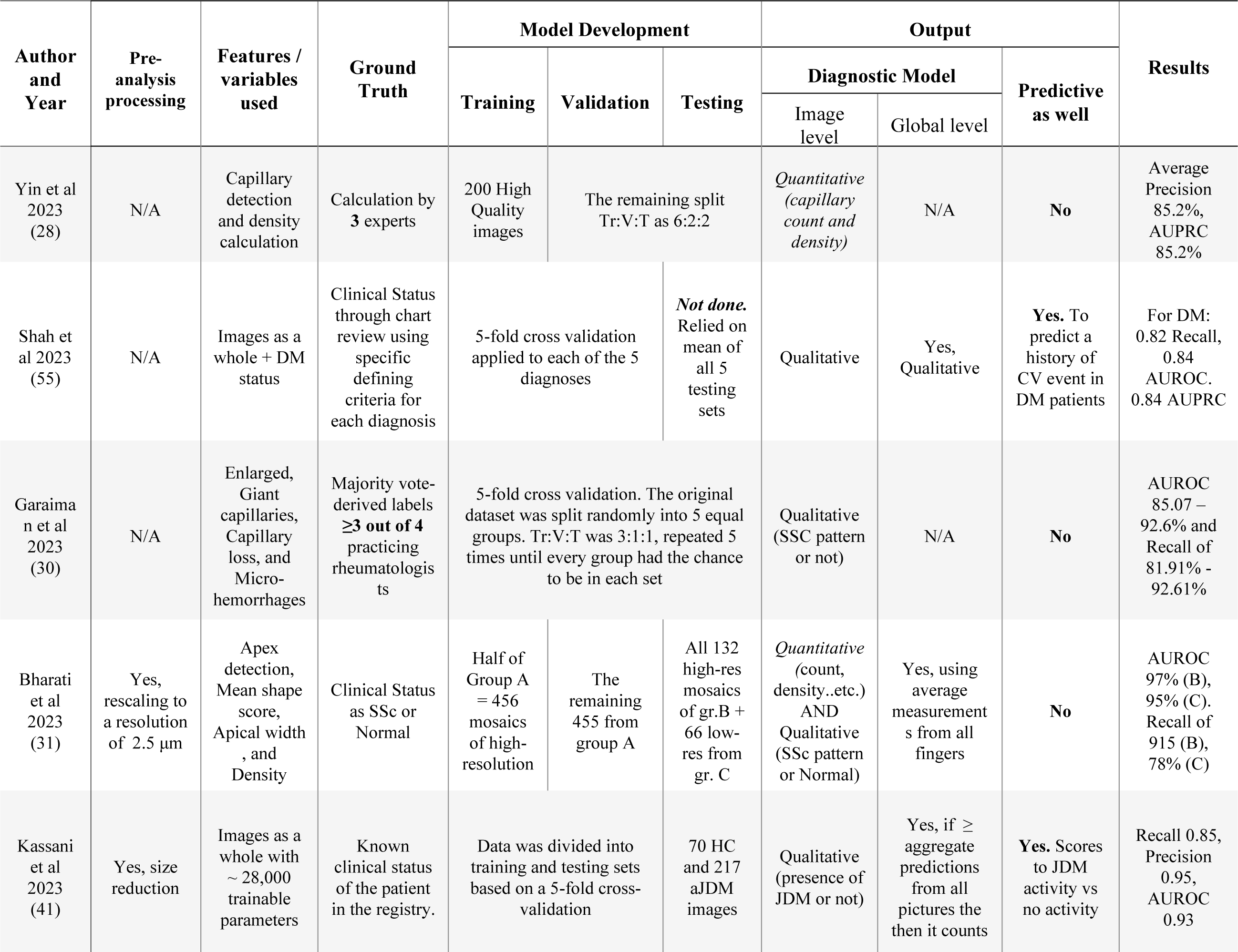
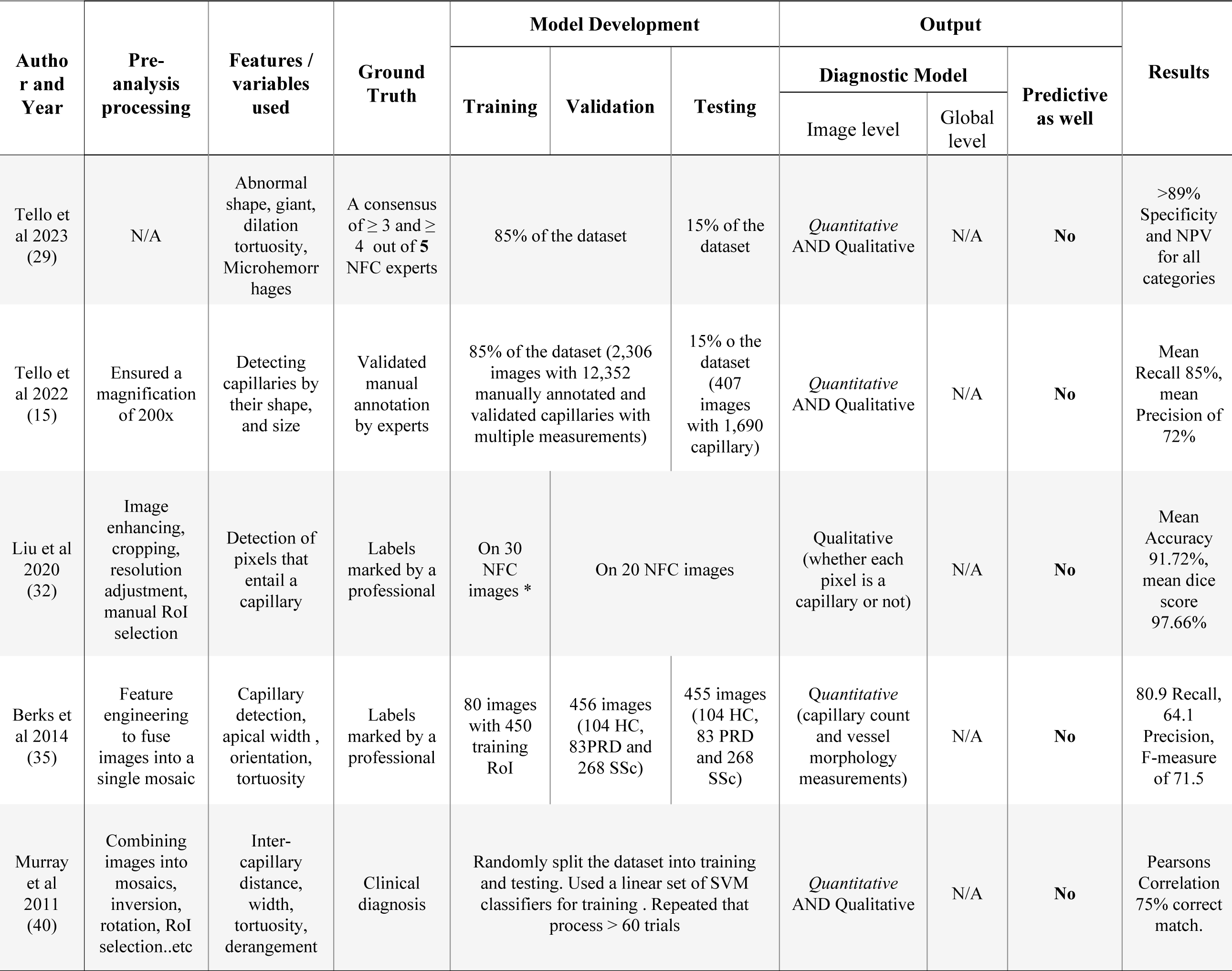
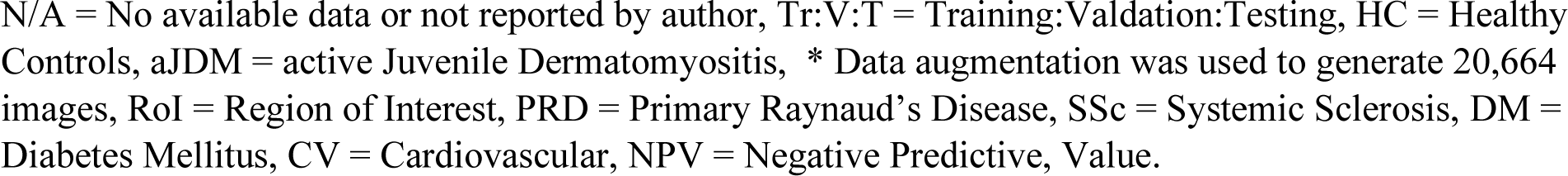
Comparing AI models across all ten studies.

The nature of tasks performed by the algorithms varied as well. The first approach employed by both Yin et al and Liu et al was a ‘segmentation-based’ object detection for capillaries (28, 32). Liu et al relied on a pixel-wise semantic segmentation to decide whether it belongs to a capillary or not (32), while Yin et al took a more global prospective to detect a capillary in the image rather than detecting it at the pixel-level (28). Four authors adopted the second approach of ‘classification’ that aimed to classify the input image into one category out of two or more categories/classes; either at the pixel, capillary, image, or global levels (30, 40, 41, 55). The remaining three authors took advantage of a multi-step mixed approach that relied on detecting certain features first, then classifying them at one or more levels into the assigned classes (15, 29, 31, 35).

#### 3.4.2 Model Input

Pre-analysis processing to optimize the model performance was carried out either manually, automatically, or semi-automatically and reported in varying detail in six studies (15, 31, 32, 35, 40, 41). It included simple tasks such as sorting the images into “acceptable” or “unusable” quality, rescaling the image resolution or size to a certain value, flipping, inversion, rotation, or brightness and noise-level adjustment. It also encompassed more complex tasks such as feature engineering to fuse multiple images of the same nailfold into a one panoramic ‘*mosaic*’, selection of the Region of Interest (RoI), marking of certain ‘landmarks’ on the image, or calculation of certain measurements to draw graphs that would later serve the model.

Eight studies reported the exact features to be extracted or calculated to be used by the model in the segmentation/classification task (15, 28–32, 35, 40). As outlined in **Table *3***, capillary detection itself appears to be the most frequently used feature, as it serves as foundation for detecting and/or calculating other metrics such as: capillary count/density, inter-capillary distance, capillary loss, apical width, enlargement, orientation, tortuosity, and derangement. On the contrary, Kassani et al and Shah et al relied on their deep learning models to find their own patterns to successfully classify NFC images, with Kassani’s model providing a visual explanation for its prediction (41, 55). **Fig. 5** depicts the process flow starting from image acquisition until the final output by the AI model is produced.

#### 3.4.3 Ground Truth Determination

Clinical status of the participant as recorded in the registry of the dataset, or charts of electronic medical records, was used as the benchmark by four authors (31, 40, 41, 55). Although this method eliminates a lot of subjectivity associated with manual labelling by experts and looks more pragmatically at the entire process through the final end-goal, it is potentially biased with documentation errors of a single expert opinion and lacks the more reliable conclusions of multiple observers. That particular downside provided grounds for the remaining authors to depend on expert annotations or labelling, whether at the capillary level or image level for ground truth determination (15, 28–30, 32, 35). They differed in the number of experts recruited (one to five experts) and in the background of the experts, whether a general NFC technician, a vascular specialist, an internal medicine physician, or a rheumatology specialist who was either a young resident or an experienced attending. Even so, the number of experts per se made a difference. For example, an odd number of 3, as implemented by Yin et al, made it easier to consider a decisive majority-vote in cases of disagreement (28). On the other hand, Tello et al who relied on 5 experts reported significant interobserver disagreement that was achieved in a non-negligible number of images (29). Authors calculated the accuracy, sensitivity and specificity of expert’s consensus and used these values as thresholds to compare the model’s performance against (30, 31, 40).

#### 3.4.4 Training, Validation and Testing

It is essential to distinguish between three different terms that entail model development: training, validation, and testing. ‘Training’ refers to the main process by which the model “learns” to perform its task, usually through multiple iterations known as epochs. ‘Validation’ usually refers to the process of internal validation, also known as *reliability testing*, in which the model’s ‘knowledge’ is tested, like a mock exam. Finally, ‘Testing’ refers to the actual assessment of the model’s performance on a set that the model was never exposed to before. Conducting model testing adds more objectivity when measuring the model performance to compare it against other models, or to the gold standard. Authors varied considerably, as shown in **Table** 3 Comparing AI models across all ten studies their approaches to splitting the dataset into Training: Validation: Testing (Tr:V:T) for model development and in their interchangeable usage of the terms validation and testing.

Three authors explicitly stated using *k-fold* cross-validation in their models (30, 41, 55). Shah et al used 5-fold cross-validation so that each random set is used 4 times for training and one time for testing by the end of the fifth round. This process was repeated for each of the 5 diagnoses (classes). They did not include additional testing using a separate dataset and they used the mean values from all 5 testing sets to be considered the final testing result (55). Garaiman et al also used a variation of 5-fold cross-validation where the original dataset was randomly divided into 5 equal-sized subsets so that Tr:V:T was done in the ratio of 3:1:1. This process was repeated 5 times until each subset had the chance to be in each set. Garaiman also had a separate dataset of 464 images that was randomly selected from the first validation subsample to test the model’s performance against manual labeling by experts (30). Finally, Kassani et al split the main dataset of 1,441 into stratified 5-fold cross-validation for both training and validation. Then a separate data set of 287 of both normal and abnormal images was used for testing the model (41).

Bharati et al separated the original dataset into 3 groups. Group A included high-resolution mosaics from 10 fingers, half of which was used for training and the other half for validation. Groups B (high-resolution mosaics from 10 fingers) and C (low-resolution mosaics from 4 fingers) were used for the final testing of the model. Thus Tr:V:T was approximately 9:9:4 (31). Yin et al first isolated 200 high-quality normal images to train the model, then the remaining 1,588 were divided into Tr:V:T as 6:2:2 (28). Conversely, Tello et al divided the dataset into 85% for training and validation, and 15% for testing in both studies (15, 29).

Liu et al divided the dataset into 60% training (30 images) and 40% testing (20 images) and compensated for the small sample by generating 20,664 images for training, from the original 30, using a data augmentation technique (32). Berks et al extracted 450 RoIs for training out of a relatively small sample of 80 mosaics. An additional set of 910 images in the dataset was split into two equally balanced sets representing all three classes; one half for internal reliability and the other for final testing (35). Finally, Murray et al split a dataset of 116 mosaics randomly into training and testing sets. Then, they utilized a set of linear support vector machine classifiers to train the model based on their labels and the associated features (40).

The methodology varied significantly between authors owing to the difference in model architecture, type of learning, balance/imbalance in the datasets, pre-processing analysis techniques, layers of networks in the model, single vs. multi-phase flow between these layers, whether feature detection was first needed before processing, and the quantitative or qualitative nature of output of the model. (15, 28–32, 35, 40, 41, 55). Additionally, authors varied in reporting the specifications of hardware and software used.

### 3.5 Models Performance

#### 3.5.1 Nature of Output

Three authors presented their model output solely in qualitative terms (30, 32, 55). Shah et al had the model classify the image out of 5 classes representing healthy status, and four different disease states (55). Gariaman’s classification of the image was binary as either diseased or non-diseased (30). Liu et al analyzed the image at the pixel level to classify whether it belongs to a distal capillary or not, so that in the end it presents a binary pixel map showing capillaries or their absence (32). In contrast, two authors reported solely quantitative reports. Berks et al reported capillary count and vessel morphology (35). Yin et al identified capillary count and density (28). Lastly, the remaining five studies combined both qualitative and quantitative results (15, 29, 31, 40, 41).

These results were majorly presented at the image level. Only three authors have also presented their results at the global, or participant-level (31, 41, 55). Bharati et al averaged measurements from multiple mosaics of multiple fingers to give a subject-level probability at a single visit (31). Alternatively, Kassani et al considered an aggregate value of ≥ 50% to be the threshold to consider a patient-level prediction successful (41).

Lastly, in addition to the main diagnostic model, only Kassani et al and Shah et al had presented a second predictive model that provided additional clinically useful information beyond merely diagnosing the presence or absence of the disease (41, 55). Kassani’s predictive model aimed to provide a score for disease activity, which translates to its future severity (41). Similarly, Shah’s model predicted a history of a complication (cardiovascular event) in patients with the disease using only the NFC images and the disease status of that particular patient (55).

#### 3.5.2 Reporting Metrics

Standardized metrics commonly used in computer vision to asses detection of objects (57) overlap with similar metrics commonly used in medicine to describe the performance of diagnostic tests. They usually entail a confusion matrix that shows the predicted against the actual positives and negatives. From those values, many indices can be calculated such as sensitivity, also known as *recall*, specificity, positive predictive value, also known as *precision*, negative predictive value, and accuracy. Certain graphs that efficiently demonstrate the effectiveness of these models are generated from these metrics and indices, such as Area Under the Receiver Operating Curve (AUROC) and Area Under the Precision Recall Curve (AUPRC). Other metrics reported included Intersection over Union (IoU), F1-scores, Pearson-Correlation, and Dice-scores. The lack of standardization of reporting presents a challenge to compare the performance of these models, which is an expected hindrance, give the novelty of this subject. It’s highly recommended that a unified approach be presented for future research so that progress can be made. However, for the purposes of this review we present below some of the significant results as reported by their respective authors.

*Recall* was reported in 7 studies, either for the final model’s ability to classify an NFC image or regarding a feature per se (15, 29–31, 35, 41, 55). It ranged from 93.87% by Garaiman et al in detecting “giant capillaries” (30) down to 70.5% by Berks et al when the model’s ability to ‘detect capillaries’ was compared with observer no.2 as the ground truth (35). *Precision* was reported in 5 studies (15, 28, 29, 35, 41). It’s highest value of 0.95 at bootstrapped confidence interval (CI) of 95% was reported by Kassani et al in classifying JDM patients from healthy controls at the image level (41). Berks et al reported the lowest precision of 51.7 in detecting capillaries when observer no.2’s labels were used as the ground truth (35). *Specificity* was reported in 5 studies (29–31, 41, 55). Bharati et al reported the highest specificity of 91% for detecting a SSc pattern in high-resolution images of group B from normal controls (31). While the lowest value of 62.2% was reported by Garaiman et al for detecting microhemorrhages (30).

*Accuracy* was reported in 4 studies only (30, 32, 35, 41). Berks et al reported the highest accuracy of 93.6% in differentiating distal from non-distal capillaries when the ground truth was determined by consensus of both expert observers (35). However, the lowest range of accuracy of 85.5% - 93.5% was reported by Garaiman et al in delineating the presence of early, active and late patterns of SSc (30). *Area Under the Curve (AUC)* was reported in 5 studies (28, 30, 31, 41, 55). The highest AUROC of 97% was reported by Bharati et al in predicting SSc patterns from normal controls in high resolution images of group B (31). Shah et al reported the lowest AUROC of 0.84 in detecting diabetes (55).

#### 3.5.3 General Descriptive Results

The time taken by the algorithm to produce the final output was not reported by most authors, except for Yin et al who reported 0.064 seconds for the capillary density calculation (28) and Garaiman et al who stated that a report can be generated for each patient with 16 images in ∼3 seconds; given that labelling 1 image takes 0.19 seconds (30). Kassani et al reported the time it takes to train 1 epoch of the model but not the final time elapsed, including any pre-analysis processing time elapsed (41).

The overall performance of all models is summarized in **Table *3***. Compared to the gold standard of manual experts labelling these models have demonstrated equally consistent performance with faster timing (28, 35), or better detection of features (30), or patterns (55). Several models outperformed other AI algorithms such as YOLO5 (28), MobileNet (41), U-Net and ResNet (32).

Some models were tested in less than ideal conditions, such as low-resolution images (31), or under different lighting conditions (28) to mimic real-life scenarios were the image acquisition process will not yield high-quality images yet the model would still be able to function effectively. Two authors reported lower performance of their models compared to experts in certain areas and they discussed possible explanations for such differences (29, 30). Finally, it is worth noting that Tello et al was the only author to do an external validation study of the previously developed algorithm at capillary.io (29).

## DISCUSSION

To answer the proposed research questions, we sought to discuss the findings of this review in four sections; given the wide variation in variables contributing to NFC analysis. Section-I lays the background by presenting the NFC technique and its significance in medicine. Section-II addresses the challenges facing the current methodology. Section-III presents the solutions AI offers to overcome said challenges. Finally, Section-IV compiles some considerations for future directions.

### Section I: NFC Technique and It’s Significance in Medicine

#### I - 1 Rationale and Premise of NFC

NFC is a simple non-invasive technique that looks at the microcirculation in the fingers. Arterial blood carrying nutritive oxygenated blood from the heart recaches one of its final destinations at the distal end of fingers/toes. NFC observes nailfolds closely through a microscope to clearly visualize the terminal capillary networks. The distal row of capillaries is seen as a convex hairpin loop that turns around to eventually form venules that carry the waste products away from tissues and back to the heart (1). This healthy microenvironment is tightly regulated and maintained so that significant changes to the capillaries array or morphology do not develop over a short period of time (3). Accordingly, chronic pathological conditions eventually distort the normal homeostasis and induce microvascular changes revealing the long-standing tissue damage (2). Examples of such systemic conditions affecting the whole body where NFC findings were correlated to disease status or progression include many rheumatological diseases such as Systemic Lupus Erythematosus (SLE) (10, 11, 58), Systemic Sclerosis (SSc) (9, 25, 59–61), Ankylosing Spondylitis (AS) (62), inflammatory diseases such as vasculitis (63–65), Idiopathic Inflammatory Myopathies such as polymyositis and Juvenile Dermatomyositis (JDM) (6, 41), inflammatory arthritis (66), dermatological such as dermatomycosis and psoriasis, and finally components of metabolic syndrome such as diabetes (55) and hypertension (32).

#### I - 2 Advantages and Significance in Medicine

NFC offers countless perks to detect such systemic manifestations. Most importantly, is the fact that it’s a non-invasive procedure that yields very valuable information that can help in the diagnosis, prognosis, staging and follow up of patients Additionally, the equipment needed is relatively cheap, and mobile, guaranteeing broad accessibility for healthcare facilities. Not only that, but the NFC technique to obtain images is by itself user friendly and easy to implement. Coupling such perks with a considerable reliability in the manual analysis of NFC images has rendered the technique the gold standard in assessing nailfold capillaries (67).

To illustrate the significance of NFC, studies of abnormal changes in the nailfold capillaries were found to be associated with skin involvement and duration of untreated JDM (68, 69). Likewise, abnormal NFC changes constitute two out of the nine scoring points to fulfill the 2013 ACR/EULAR classification criteria of SSc (70) and is important in stratifying SSc patients into early, active and late (71–73).

It is also used to differentiate between types of Raynaud’s *Syndrome* (RS). RS is an episodic color change in the fingers with or without pain in response to cold. It can present as a primary benign condition with no evidence of an underlying disease, known as Primary Raynaud’s Syndrome OR Raynaud’s *disease* (PRD). Or it can be a secondary symptom associated with other medical conditions, like SSc (56), one of the most common causes of Secondary Raynaud Syndrome or Raynaud’s *phenomenon* (SRP). Trombetta et al found that a quantitative change in the capillary diameter is predictive of progression of PRD into SRP (17). Dolijanovic et al followed a cohort of 250 PRD patients over six years and found that most participants had normal findings with only 10 out of all subjects (4%) would show SSc pattern 6 months before expressing a particular disease (74). Although, this study concluded a considerable lack of reliability in predicting future progression to certain diseases from such non-specific capillary changes in that study population, it did highlight an important finding, and that is if a SSc pattern was found, it would highly correlate to future development of SSc with high specificity and precision.

#### I - 3 Preprocedural Preparation

Before the procedure, participants are prepared by avoiding caffein, smoking, stress, and cosmetic procedures on fingers/toes for about 3 weeks before the test day. Then, they are seated in an upright position where the heart is at the level of the nailfold, in a quiet room with a stable temperature of >20 °C for about 15-20 min to provide heat adaptation and endure mental comfort. Next, the nails are cleaned, and an immersion oil is applied to improve visibility and translucency by reducing diffuse reflections (75).

The apparatus is then set after deciding the following key aspects. The model and type of the capillaroscopy device is determined and whether it will be fixed or hand-held. Then the magnification is set depending on the camera resolution, its angle and physical distance from the nailfold (contact vs on-contact). Note that calibration to account for such distance might be required; either manually or automatically. Next, the finger/toe to be examined is placed and the light source, its intensity and angle are adjusted for optimal brightness and least reflections.

#### I – 4 NFC Image Acquisition

Traditional NFC relies on a camera to capture photos of the nailfold. NVC is an updated alternative that records a video instead, then extracts screenshots of frames with good capillary visibility. Some authors like Murray et al described a software that allowed high-magnification panoramic mosaics to be constructed from a video without movement artifacts (40). After images are obtained, they might be manipulated and *pre-processed* to varying degrees, either manually, or using additional software, before they are finally fed into the AI model.

### Section II: Current Challenges Facing the Manual Approach

The current golden standard encounters several significant challenges, owing to the sheer variance in variables and circumstances required to obtain NFC images and analyze them. We also comment on how certain authors have approached that area and consider how AI can potentially overcome these obstacles.

#### II – 1. Lack of standardization of the procedure (technique homogenization)

##### II – 1.1 Preprocedural Matters

Starting with patient selection, most studies were unsuccessful in reporting significant demographics related to the selected sample. At the top of the list, differences in skin tone and pigmentation. This vital factor could point towards a potential bias in acquiring images of fair-skinned participants only or in the reliability and validation of the technique itself in dark-skinned patients, propagating error to be carried forward when training and developing the Machine Learning (ML) or Deep Learning (DL) models. With regards to the detailed technicalities of patient preparation before image acquisition, only Garaiman et al intentionally stated referring to the international Delphi consensus described by Ingegnoli et al (75); taking a step forward towards unifying the pre-procedural setting.

Consider the two following apparatuses that were described by some authors: a contact hand-held Optilia capillaroscope versus a non-contact fixed Dino-Lite microscope. The first setup permits a broader-angle adjustment and higher zoom to improve visualization, but it runs the risk of unsteady operator hands, and thus image blur due to movement artifacts. On the contrary, the second setup eliminates movement artifacts and can provide a more panoramic view of the entire nailfold, yet it requires higher magnifications and resolution to compensate for the increased distance between the camera lens and the nailfold surface. Low magnifications such as 50x taken by dermatoscopes can provide a broader view of the nailfold, but are not detailed enough to reliably discern capillaries when compared to the more ideally desired magnification of 200x (76).

A capillary’s morphology varies between hands and feet, finger to finger, and even central to peripheral nailfold sectors (77). According to Cutolo et al, the gold standard technique is to capture at least two adjacent fields of 1mm in the center of the nailfold at 200x (76). Dinsdale et al reported that examination of all 8 fingers is needed, excluding thumbs. Otherwise missing some abnormalities will reduce sensitivity(78). Conversely, Murray et al reported high model performance using only the 4^th^ (ring) finger in the non-dominant hand (40).

##### II – 1.2 Procedural Considerations

Surprisingly, the duration of the whole process from image acquisition till final output was not reported by many authors. The breakdown of training time, pre-processing time and execution time is also recommended to be reported in future studies to calculate the net time elapsed. Considering how much time pre-analysis processing consumes, it’s variation from single simple tasks to complex multi-step editing might necessitate user expertise and training. It is of vital importance that authors report the software used, its version, specifications and tasks performed so that appropriate comparisons can be concluded. For instance, shah et al fed the DL model with images only, to receive the final output (55). On the contrary, Tama et al (79) and Doshi et al (80) adopted complex multi-step approaches such as binarization, skeleton extraction and segmentation, and enhancement operations with alpha-trimmed filter to address non-uniform lighting combined with an iterative rule-based skeletonization procedures.

Additionally, Other aspects are not agreed on unanimously, yet. For example, AlGindya et al reported that according to the literature, the green channel of an RGB image shows high contrast between capillaries and the background (81), and therefore used the green filtered images for input. Contrarywise, Liu et al relied on the greyscale because they found that there were no significant differences between the greyscale and the green channel image (32). Another example reported by Liu et al, is that they claimed vertical flipping of an image might change its semantic information in the object hierarchies, in contrast to the horizontal flipping (82).

Finally, we emphasize the need to report the specifications of hardware used, in addition to the software. Such factors might be limiting in terms of computational costs, timing, and even critical decisions such as feeding multiple measurements as input versus relying on a single average value, hence determining the level of processing, pixel-wise, capillary level, or image level.

#### II – 2. Image Ambiguity

The ambiguity in images is usually a result of its poor quality and/or presence of artifacts that could be due to a wide range of factors. Camera-related factors include low contrast/resolution images (mostly in low-cost devices), high image noise, lighting issues such as reflections on the oil, non-uniform lighting, and extremes of brightness. It could be due to physical factors such as air bubbles in the immersion oil, dust on lenses, dirt on/in fingernails, or blurring due to movement of patient fingers during imaging or examiner’s hand if it’s a hand-held device. Lastly, such variance may be disease-related such as too much fibrosis as in the late stages of SSc, or due to presence of non-delineable structures. Poor reporting and lack of standardization in tolerance thresholds to all these procedural parameters will result in increased image heterogeneity and accordingly, higher interobserver variability that will eventually be transferred to the ML model.

#### II – 3. The Subjective Nature of Analyses by Human Experts

Manual analysis of NFC images, whether by clinicians or trained personnel, exhibits considerable subjectivity that massively influences the gold-standard technique as well as the input to ML/DL models, and therefore their predictions. Firstly, *operator bias* is demonstrated through their reliance on intuition to select fields, capillaries and in classifying them instead of examining all capillaries in each image. It also manifests as rough estimates of features instead of relying on accurate measurements and indices. Finally, numerous cognitive biases could develop if analysis is done, for example, right after reviewing a patient chart. Other expected shortcomings due to human operators include the need for training to raise expertise (23) in addition to user, owing to the time consuming tasks to identify, label structures, categorize them and calculate indices; especially when eight fingers are examined to maintain a high sensitivity (78).

Inter-individual variability is yet another significant aspect due to the multi-variable nature of the task. Despite testing its reliability, high interobserver variability still poses a considerable bias especially when images get less clear/more ambiguous (67, 83). Saez et al in the GEAS survey found considerable heterogeneity between capillaroscopy experts, particularly when considering morphological differences and not just categorical normal versus abnormal patterns (84). Similarly, Garaiman et al reported high agreements especially regarding giant capillaries and microhemorrhages, as well as regarding assessment of patterns (e.g. SSc Pattern), but not very much regarding capillary loss and enlargement (30). Such high inter-rater variance presents a challenge, per se, when the performance of the algorithm is judged against experts as the decision of which expert to be regarded as the benchmark is made. Thus authors like Tello et al 2023 (85) and Garaiman et al (30) reported an acceptable decent performance by the algorithm despite most experts performing better than the algorithm. Furthermore, the agreement is not only low with regards to the grading of each image, but also in selecting which areas to be evaluated (35). That was the basis for a more standardized criteria called the ‘fast track algorithm’ that was developed to help ease and standardize the grading process (60, 72, 86, 87).

#### II – 4. Lack of Agreement on Which Features to be Extracted?

Current practice only looks at the distal row of nailfold capillaries. Some general features of a normal nailfold include a transparent skin with clearly visible capillaries, absent pericapillary oedema, visible subpapillary venous plexus (in up to 30% of healthy people), and similar-looking capillaries that are regular in arrangement, mostly straight and perpendicular to the nailfold (88).

Most of the features considered for analysis are qualitative and they can be a singular feature such as a U-shaped hairpin-convexity constituting the normal capillary morphology. Or it could be the absence of singular features such as tortuosity, ramifications, neo-angiogenesis, 3-point crossing, non-convex tip, and hemorrhages. (89, 90).

An additional way of analysis is through a collection of certain features, known as a pattern, that defines a certain abnormal condition from the normal population. For example, Raynaud’s Syndrome is an episodic color change in the fingers with or without pain in response to cold. Raynaud’s syndrome can present as a primary condition with no evidence of an underlying disease, known as Primary Raynaud’s Syndrome OR Raynaud’s disease (PRD). Or it can be a secondary symptom associated with other medical conditions, like SSc (56), one of the most common causes of Secondary Raynaud Syndrome or Raynaud’s phenomenon (SRP). To distinguish PRD from SRP, Mannarino et al (91) used the following pattern: altered arrangement of capillary loop, decrease in the number of capillaries, and abnormal ramifications. Diagnosing SRP is significant because it is considered a reliable early parameter for diagnosing early SSc that has been clinically validated. One important way of doing that distinction, according to Bharati et al is that if PRD are negative clinically and serologically, they are likely normal (31).

Another very common and significant example relates to SSc. It is a potentially lethal autoimmune disease characterized by 3 hallmarks: Microangiopathy, production of disease-specific autoantibodies and deposition of extracellular matrix proteins resulting in tissue fibrosis. (70, 92) The microangiopathy usually manifests as low capillary density, high capillary dimensions (dilation or giant), and abnormal morphology and hemorrhage (73, 93, 94). Smith et al defined SSc pattern as very low capillary density (<3 capilaries.mm), or the presence of giant capillaries (72). Murray et al added high tortuosity and derangement; that is disorganization in the overall direction of capillaries (40).

The same disease, like SSc, could show different patterns across different stages of the same disease. that is typically used to help monitor improvement or disease progression on follow up. For example, early, active and late/severe patterns of SSc have been identified and validated in clinical studies, commonly the one described by Cutolo et al and standardized by Smith et al. (17, 25, 95, 96). The importance of recognizing such patterns helps monitor disease progression or improvement on follow up. This finding presents an opportunity for early intervention before severe organ involvement could occur, or potentially predicting future disease status. Finally, Other less understood non-specific set of features are as known as disease-associated changes (15, 30, 85)

A few quantitative features have also been described such as avascular areas defined as distance between 2 loops > 500 μm, that normally should be absent (88). Interestingly, it was demonstrated that calculation of the mean score of such capillary loss could be reliably reduced from 32 NVC images (four fields per finger for eight fingers of the patient analyzed) to eight NVC images (one field per finger for eight fingers of the patient analyzed). This finding can save valuable patient time as well computational costs, stressing on the significance of disclosing all aspects of the process to help deduce such conclusions (97).

Another commonly calculated feature is capillary density (normally = 9-13 in 1 linear mm). Density is related to tissue perfusion and microvascular function. The less normal conditions are the lower the density. Finally, many measurements can be calculated for each capillary, including: arterial limb diameter, venous limb diameter, apical loop diameter (7) (normally < 20 μm (25)) and total width. From these calculations, two important descriptions emerge, namely a dilated capillary (typically 20-50 μm), or a giant/mega capillary (typically >50 μm) (25) that are normally absent in a healthy individual.

#### II – 5. Lack of Standardization of such Features

##### II – 5.1 Defining a Parameter or Pattern

Unfortunately, the aforementioned features lack unanimous agreement on a validated and clinically relevant definition. To demonstrate, let’s consider the capillary density. Generally speaking, Neubauer et al described a normal density as 9-13 per linear mm (88), while Cutolo et al relied on capillary loss as <7 capillaries per mm (98). Alternatively, in et al didn’t rely on a specific criteria and they rather compared their model’s performance to that of the experts count in normal healthy volunteers (28). On the other hand, Bharati et al were more specific in their definition as they counted all capillaries from left-most to right-most within each mosaic image of the entire nailfold and divided that number by the distance in mm between the same left-most to right-most (31).

Another variation lies in the approach to detecting a terminal capillary loop/apex in the distal line of capillaries to differentiate it from other detected vascular structures. For instance, some authors rely on the direct observation method compared to the 90° degrees method described by Hosftee et al (67) and used by Yin et al (28). Alternatively, Karbalaie et al developed the semi-automated Elliptic Broken Line (EBL) method (99). Recently, Bharati et al used the fully automated deep learning model U-Nets for apex candidate generation, followed by ResNet34 for candidate classification (31).

The difficulties due to lack of such standardization is not unclear, as expressed by Cutolo et al, regarding the reliability of simple capillaroscopic definitions in describing capillary morphology in rheumatic diseases (89). In an attempt to overcome these differences some authors proposed potential standards to be followed whether in defining a single parameter or a pattern. For instance, Jones et al proposed a taxonomy for the morphology of nailfold capillaries (38). Neu-bauer Geryk et al described a possible standardized technique with proposed normal values to define a normal for healthy subjects (88). Smith et also developed well-defined criteria that was adopted by the EULAR society to describe normal (<20 μm), dilated (20-50 μm), and (giant/mega >50 μm) capillaries (25). Smith et al and Cutolo et al attempted to standardize the defining criteria for SRP in SSc and how it differs from PRD (25, 72, 73). Cutolo et al also initiated a similar proposition for a different disease like SLE (11).

##### II – 5.2 Clinical Relevance and Validation of Features

Surely, studying these features without any clinical relevance considerations or reliability testing will render the entire process purely academic and hinder its progression to a practically applicable tool. Such studies will emphasize the ‘more significant’ features so that they get assigned a higher weight. As an example, Trombetta et al found that a capillary diameter of > 30 μm is an independent predictor for the progression of PRD patients to SRP (17). Similarly, capillary density was found to be a significant quantitative parameter in studies of conditions like diabetes (100), connective tissue diseases, pulmonary hypertension in SSc patients, and chronic kidney disease (99) that would relate to a particular aspect of these diseases. Understanding the implications of this matter, the SCLEROCAP study by Boulon et al was conducted to assess the reliability of Maricq and Cutolo’s classification of capillaroscopic features that can stage SSc patients, or rather predict their status (101). Accordingly, efforts to propagate and disseminate the validation of these features should be encouraged to guide future research in towards a more patient-oriented direction, whether these features were relating to the study of condition at hand, or other unknown conditions. Moreover, certain features might be deliberately, and safely, ignored, like what Bharati et al did. They disregarded microhemorrhages from being input into the model since they saw the difficulty to define it with lack of its relevance to diagnosing SSc (31).

For that reason, authors like Tullo et al (29) decided to detect a feature such as *tortuosity*, even though it had no current validating studies, to understand possible ways it could prove relevant. Typically, it has been looked at with a quantitative approach, calculating the percentage of tortuous capillaries to discern normal healthy individuals (<5%) from other conditions that could potentially manifest at certain higher cutoffs such as 40% or 70% (102).

##### II – 5.3 Decisions in Conflicting Matters

Defining hierarchical principles in labelling a capillary is an important step that needs to be addressed especially after these labels will be used to train ML models. For example, Tello et al (103) prioritized classifying a capillary as being ‘giant’ over ‘tortuous’, if both features were present in the same capillary. Additionally, Garaiman et al noticed sub-optimal consistency in NFC image labels owing to the simple fact the different attending physicians have analyzed images of the same patient at different time points, thus affecting the ground truth of the developed model (30). Another decision would arise if the feature at hand is not clearly visualized. For example, Tello et al relied on the width of limbs as an acceptable alternative (in case the apical diameter wasn’t clear enough) to detect dilated/giant capillaries(29).

Furthermore, when experts, and henceforth, ML models, aim to assign patient-level diagnoses, the approaches differ. For example, some experts would label a participant as abnormal if ‘any’ NFC image from anyone of the finger is labelled as abnormal. Conversely, Bharati et al averaged measurements from all participants to achieve global-level labelling (31), while Kassani et al considered a participant diseased if the aggregate from all pictures was ≥ 50% (41).

### Section III: How AI Is Solving These Challenges

#### III – 1 Evolution of AI beyond simple automation

Artificial Intelligence (AI) was first described as a term by John McCarthy in 1956 as the science and engineering of making intelligent machines. Despite the difficulty of defining exactly the aspects of human thinking, rationalization and decision-making processes, AI aims to simulate one or more of these complex processes, at least partly. Automation of any process relies on algorithms which are mathematical operations or code, that can be simple or a multi-step series, that are fed a particular type of input to process in a precisely defined manner and give a certain output. If it was given any new or unusual input that it’s not programmed to process, it will fail to deal with it. AI, however, is more than a simple automated code, that performs ‘smart’ functions, in an attempt to mimic human reasoning. ML, which is a subtype of AI, takes it a step further, by learning how to deal with new information on its own without much prior programming, exactly like a human child learning. DL is a further subtype of ML that tries to learn entirely on its own by interacting with its environment in certain ways and developing a ‘brain’ of its own that is a black box to its developer.

A common way of teaching these machines how to learn is the supervised approach through input-output pairs. In our case, this means showing the model a normal NFC image and telling it that it is normal or showing it a particular feature and telling (labelling) it what it is. Another would be to let the model figure out differences in input on its own by feeding it only the images, known as unsupervised. Other methods employ a mixture of both include semi-supervised and reinforcement learning whether using a human agent in the loop or not. Although ML is a smart enough machine that can do incredibly difficult tasks, DL models seem to look at things through a ‘fresh set of eyes’ that offers an unparalleled and novel approach to analyses.

#### III – 2 Superiority of AI

Overcoming the challenges that come with NFC image ambiguity (as discussed in section II-2) had posed a huge obstacle for experts to interpret these images. AI models can detect minute structures that are invisible to the naked eye and find relations between an incredible number of parameters within a very short time frame. Such qualities allow it to circumvent such heterogeneities, or even edit them before analyses in a fully automated, and possibly real-time, manner.

AI has also eliminated a lot of (not all) the errors attributed to human operation. Provided with decent hardware, an AI model can compute an insanely massive number of parameters at the same time to produce a consistent more objective output within seconds, if not less. Its training is less exhaustive and consuming compared to human training, and it would be able to analyze so much more with less resources. Such capabilities can potentially present a more objective output that relies on certain criteria, compared to human experts. As an example of how AI models can reduce operator bias, can be demonstrated as many experts usually rely on their subjective intuition to select a few capillaries within a sector of an NFC image of a single finger to classify it rough manner. An AI model, will instead look at all the capillaries in the entire nailfold of all fingers, take precise measurements and draw indices related to all of them, extract much more features and issue a more comprehensive and thorough judgment in a fraction of the time taken by experts. Not only that, but its output wouldn’t be influenced by reading patient’s charts, as might happen with experts.

Additionally, future predictions about the potential risk of developing a certain condition can be drawn. Current models have demonstrated the potential to fill the gap in identifying predictive features correlating to actual diseases progression (33).

### Section IV: AI Considerations and Future Directions

#### IV -1 Current Limitations and Challenges

Thorough documentation and comprehensive reporting of the entire process, from patient selection and pre-procedural preparation to image acquisition and analyses cannot be overstated. It is essential to recognize that as long as humans exist, their biases will too. The question therefore becomes how to minimize propagating these biases to AI. For example, the scarcity of datasets in dark skinned population will render these AI models inefficient in performing the same task compared light-skinned populations.

Similarly, errors and biases in the preprocessing, as pointed out by Murray et al may shift towards a particular parameter more than others, and eventually skew the mode towards over/under estimations (40). Moreover, standardizing the measurement units’ whether real or arbitrary will close the gap towards a more objective feature extraction and therefore, output. An additional drawback is the limited open-access datasets that newer models can use for training.

Condition-specific factors may also limit the predictive models’ accuracy. For example, Kassani et al reported difficulty in predicting JDM disease activity due to difficulty in measuring disease quiescence (and limitations in doing so using the manual DAS itself, as it fails to sensitively capture disease activity compared to other recent biomarkers) and the fact the capillary damage may persist with inactive disease(41). That example enforces the fact that biases are carried forward with the AI, and that the burden of providing such answers falls on the developer, not the AI model. That is not to say that AI models don’t share the burden of ethical responsibility to be explainable, but that it highlights the usually overlooked human bias.

#### IV-2 The Future of AI in NFC

AI models can be further enhanced by feeding them with more information that just an NFC image, such as a patient’s electronic medical record, similar to what Sheh et al did (55). Impressive results already established when NFC was fused with other modalities such as ultrasound (104, 105), doppler sonography (106), laser scanning microscopy(107), and optoacoustic imaging (108, 109).

## CONCLUSION

AI models have demonstrated a truly remarkable potential as a clinical decision-supportive tool. Owing to the novel nature of this technology, it is of the utmost significance for authors to report future NFC-related studies in the most comprehensive way possible, particularly population demographics and execution times. That is to overcome propagating human biases to AI models, standardize the NFC methodology and reporting metrics to allow for comparisons and conclusions to be made. ML and DL models succeeded in producing a fully automated and objective quantitative output that will form the basis for future prediction and patient-reported outcomes research. Fusion of NFC with other technologies like doppler laser and optoacoustic imaging to enhance the extraction of features and the precision of measured values will further increase the sensitivity and specificity of such tools to be very efficient in daily clinical practice. Future studies should focus on the deployment and provision of full functionality for these types of applications using explainable AI. Quality assessment standards and ethical considerations still present a big challenge in reporting and in testing the safety of these techniques. Finally, with more external validation studies across multiple different settings in different populations these tools will revolutionize diagnostics through NFC image interpretation.

## Data Availability

The data extraction tables, figures, and supplementary documents will be uploaded alongside the main manuscript.

## OTHER INFORMATION

### FUNFING AND CONFLICT OF INTERESET

The authors of this review declare no potential conflicts of interest regarding research, authorship, or publication. This review was not supported by any source of financial aid.

## ACKNOWLEDGMENTS

Figure 1 was created using MS Word

Figures 2, 3 and 4 were created using MS Excel

Figure 5 were created using MS Paint

## LEGENDS

Fig. 5 A summary of the whole process of NFC and AI analysis.

**S.1** Shows the risk of bias and quality assessment of all included studies using QUADAS 2.

## Notes

**Financial Disclosure Statement:** None of the authors have any conflicts of interest.

### Competing Interest Statement

The authors have declared no competing interest.

### Funding Statement

The author(s) received no specific funding for this work.

